# HIV Prevalence, Virological Suppression, and Consistent Condom Use among Social Venue-Going Men in Zimbabwe: Insights from the 2022 Priorities for Local AIDS Control Efforts (PLACE) Surveys

**DOI:** 10.1101/2025.06.18.25329843

**Authors:** Galven Maringwa, Sungai T. Chabata, Fortunate Machingura, Jaspar Maguma, Memory Makamba, Tariro Chinozvina, Samson Chikura, Leslie Nyoni, Madonna Mlambo, Edward Matsikire, Amon Mpofu, Raymond Yekeye, Benard Madzima, Owen Mugurungi, Brian Rice, Sharon Weir, James R. Hargreaves, Elizabeth Fearon, Frances M. Cowan

## Abstract

**Introduction:** Men attending social venues face barriers to accessing HIV prevention and care services. These venues—such as bars, guesthouses, nightclubs, and transport hubs—facilitate new sexual partnerships but lack cohesive social networks, making it challenging to design and implement effective HIV prevention strategies. Men who attend social venues are more likely to pay for sex, potentially increasing their risk of acquiring or transmitting HIV. However, data on how HIV-related behaviours and service engagement differ between men who do and do not pay for sex among those attending venues remain limited. This study examines whether men who pay for sex have higher rates of HIV prevalence, lower rates of virological suppression, and poor HIV-prevention-related behaviours compared to those who do not.

**Methods:** Using the Priorities for Local AIDS Control Efforts (PLACE) methodology, we collected cross-sectional data from April to December 2022 across 190 venues in four cities and towns in Zimbabwe. Participants underwent finger-prick HIV testing; those testing positive provided dried blood spots (DBS) for viral load measurement. We also collected sexual behaviour data, including condom use. We applied survey weights and used weighted Poisson regression models with robust standard errors to investigate factors associated with HIV status, virological suppression, and condom use among venue-going men, treating paying for sex as the primary exposure variable. All reported percentages are weighted.

**Results:** Among venue-going 2,827 men, 984 (40.1%) reported paying for sex in the past 12 months, and 531 (15.1%) reported consistent condom use in the past month. Overall, HIV prevalence was 10.7%. Among men living with HIV, virological suppression was 67.9%. In adjusted analyses, there were no significant associations between paying for sex and HIV status (adjusted prevalence ratio (aPR) = 0.89, 95% CI: 0.52–1.55), self-reported consistent condom use in the past month (aPR = 0.87, 95% CI: 0.57–1.34), or rates of virological suppression among men living with HIV (aPR = 1.03, 95% CI: 0.75–1.42)

**Conclusion:** Findings indicate substantial HIV risk and suboptimal prevention and treatment engagement among men frequenting social venues, irrespective of paying for sex. Therefore, targeted interventions are needed for both paying and non-paying men.

## Introduction

Though there has been broad progress in coverage of HIV treatment and expanded access to prevention services, HIV/AIDS remains a serious global health problem, particularly in sub- Saharan Africa, which harbours nearly two-thirds (65.0%) of all infections(1). Men who patronize venues and pay for sex risk HIV and sexually transmitted diseases (STIs) and can infect their partners(2, 3). According to Zimbabwe’s 2015 Demographic and Health Survey (ZDHS), 4.0% of men aged 15–54 reported paying for sex in the past 12 months(4). In 2022, clients of FSWs accounted for an estimated 10.0% of new HIV diagnoses globally(1).

Despite extensive research on HIV epidemiology and service uptake among female sex workers (FSWs), data on men who have sex with FSWs remain limited. This gap includes information about their patterns of condom use, pre-exposure prophylaxis (PrEP) uptake, voluntary medical male circumcision (VMMC) status if HIV negative, and engagement with treatment if living with HIV (LHIV). Although effective treatment adherence leads to virological suppression and prevents sexual transmission(5), men generally have poorer outcomes than women across the HIV care continuum, from HIV status awareness to optimal use of ART and virological suppression, and are therefore characterised as ‘left behind’ in HIV response efforts(6).

As a result, these men are often underserved in terms of HIV services because they are difficult to target as a coherent population, contributing to higher rates of HIV transmission. Social venue-going men exhibit high-risk behaviours associated with HIV(7–9). Among men recruited at social venues in South Africa, those who were 25 years or older, had not completed high school, did not use condoms at first sex, consumed alcohol or substances, or were not medically circumcised were more likely to have a positive HIV status compared to men without these characteristics(10). Social venue-going men also exhibit low uptake of HIV prevention services, including consistent condom use(9). Consistent condom use offers dual protection against both STIs, including HIV, and unwanted pregnancies, and it is highly effective(11, 12). Condom use with heterosexual partners is influenced by a range of socio-demographic, behavioural, psychosocial, relationship, contextual, and structural factors(13). Factors associated with inconsistent condom use include age at sex debut, illicit drug use(14), marital status or partner type(15–20), and perceived high self-efficacy for condom use(21).

To inform prevention and treatment programming, we assessed HIV prevalence, virological suppression, and HIV-related behaviours among men at social venues in four Zimbabwean cities and towns. We compared the risks of HIV acquisition and transmission, care needs, and engagement with HIV prevention and care services among men who reported paying for sex and attending social venues in the past 12 months with those who did not report paying for sex during this period.

## Methods

### Study design and population

Between April and December 2022, we applied the PLACE methodology to collect data from venue patrons and workers in four cities and towns across four different provinces in Zimbabwe (Figure 1). PLACE aims to generate detailed, locally relevant data to guide HIV prevention and care by mapping social venues where individuals meet new sexual partners, using a systematic, multi-phase approach that includes stakeholder engagement, community informant surveys, venue informant surveys, and biobehavioural surveys (BBS)(22–30). In PhaseD1, we worked with stakeholders (e.g., provincial medical directors, local health officials, HIV response partners, community advisory boards, monitoring and evaluation directors, and the National AIDS Council) to identify Priority Prevention Areas (PPAs). PPAs are geographic locations, venues, or subpopulations where the risk of HIV transmission and acquisition is highest, and where targeted interventions can have the greatest population-level impact by reducing new infections and improving engagement in care. In Phase□2, we generated lists of social venues (e.g., bars, guesthouses, nightclubs, and transportation hubs) by asking knowledgeable community informants (e.g., bartenders, police, taxi drivers, street vendors, and peer educators) to name all the venues they knew (community informant survey). In Phase□3, we visited all previously identified venues and classified them as operational, not found, temporarily closed, permanently closed, or duplicates (venue informant survey). Operational venues were profiled for operating hours and busy times, activities and amenities, the availability of onlJsite prevention services such as HIV testing, peer education, condom distribution, and estimated attendee populations. Finally, in Phase 4, we conducted the BBS at a random sample of these venues on busy nights, interviewing patrons and staff and offering HIV testing.

**Figure 1:**
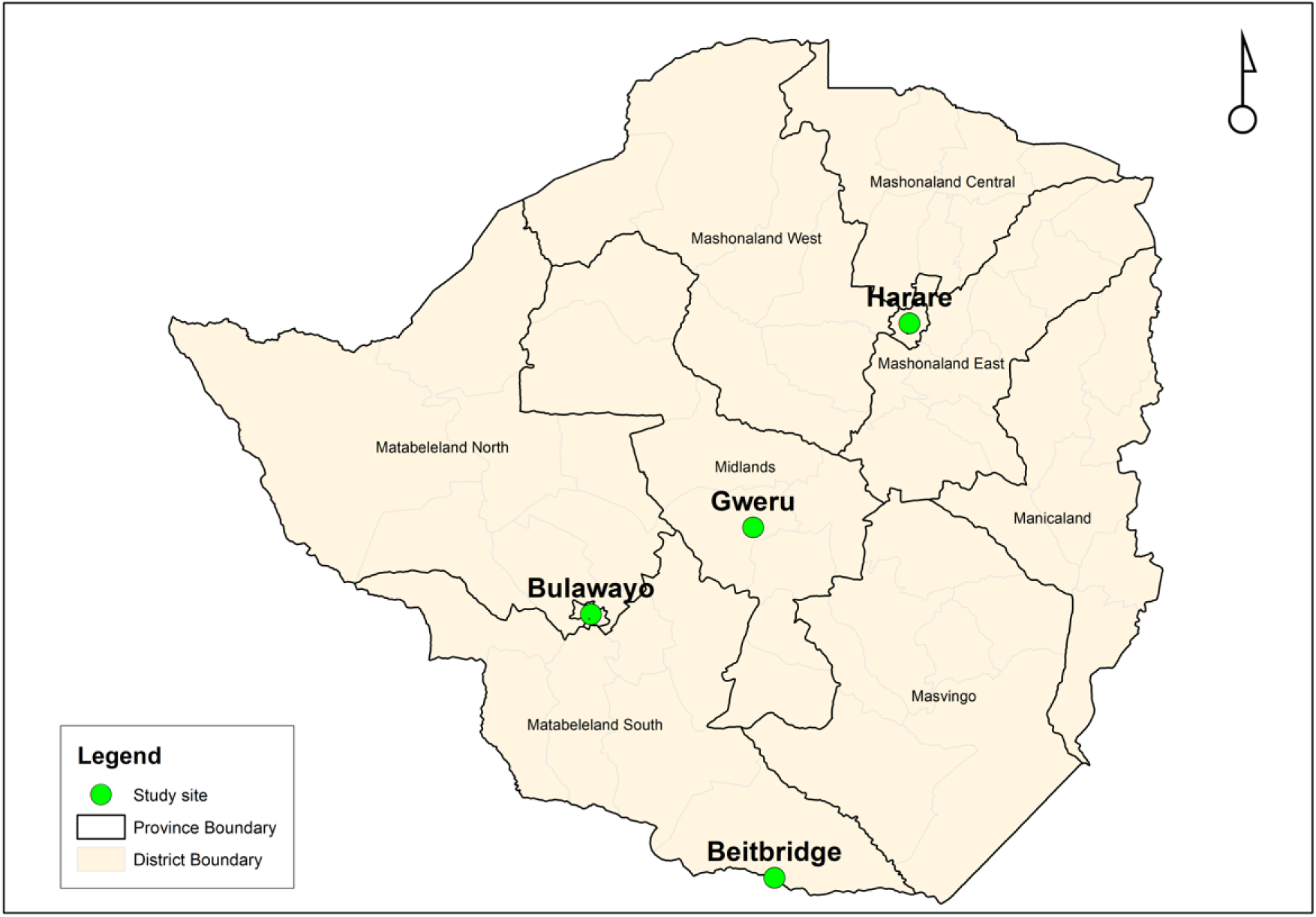
Map of Zimbabwe showing PLACE study cities and towns

In Bulawayo, Zimbabwe’s second-largest city, stakeholders purposively chose one of three PPAs because, unlike other PPAs, it encompassed each identified venue type and the Central Business District. Among 198 venues identified in this PPA, 52 were randomly selected via stratified sampling. In Gweru, a mid-sized city, 190 venues were identified and 30 were randomly selected for the BBS. In Beitbridge, a key border town adjacent to South Africa, characterised by significant cross-border traffic, 143 venues were identified and 30 were selected for the BBS. In Harare, the capital and largest city, 2,456 venues were mapped across 196 districts, and three districts were purposively selected for the BBS based on District Technical Working Group recommendations. From these districts, 78 venues were chosen through stratified random sampling. At each selected venue, BBS participants included male and female patrons and workers aged 16 years or older who were present during the evening hours on Friday, Saturday, and Sunday. We recruited all workers in venues with fewer than 10 workers.

### Data collection

The number of individuals interviewed and tested for HIV at each venue was proportional to the expected attendance provided by venue informants during the venue informant surveys. All on-site participants provided informed consent by marking an ’X’ on the consent form, while witnesses signed the forms. Participants then completed an interviewer-administered questionnaire. No data on refusals was collected. All men gave a finger-prick blood sample for HIV testing. To allay concerns regarding privacy and confidentiality, HIV tests were performed privately and anonymously at the venues by trained research nurses with valid Good Clinical Practice certificates, and participants received their results immediately after testing. Each participant received USD 2 for their time. Men newly diagnosed with HIV were referred to key population (KP) clinics or initiation sites within their districts for antiretroviral therapy (ART). They received a referral slip to present at these sites, and research teams coordinated with facility personnel to ensure prompt ART initiation. Viral load tests were conducted on HIV-positive individuals using Dried Blood Samples (DBS), which were stored and shipped in zip lock bags with desiccation packets(31). Participants were later contacted to obtain their viral load results at KP clinics, with participants indicating the specific clinics where they wanted to access their results. Questionnaire data included demographic characteristics, venue going behaviour, sexual behaviour, condom use, and uptake of HIV prevention and care services. Other variables included the Alcohol Use Disorders Identification Test (AUDIT), a screening tool used to assess potential hazardous alcohol use(32), and the Shona Symptom Questionnaire-14 (SSQ-14) to assess psychological distress and symptoms related to common mental disorders (CMD)(33).

### Data management

All HIV results and questionnaire responses were entered into a database and linked using unique participant identifiers. Data from all sites were pooled for analysis. Pooling ensured greater statistical power due to increased sample size and resource optimization by conducting a single analysis compared to site-specific analysis.

### Measures

HIV testing was conducted to determine the prevalence of the infection. Viral load (VL) testing was performed using the Abbott RealTime HIV-1 viral load assay (Abbott Molecular, Des Plaines, IL, USA) to test the two whole blood spots extracted from each card and eluted in RNA lysis buffer (Promega Corporation, Madison, WI, USA). Undetectable viral load (VL), that is, virological suppression, was defined as a VL of <1000 copies per millilitre. For mental health (SSQ-14), a score of 9 and above indicated symptoms of CMD, and for alcohol use (AUDIT), a score of 8 or more was associated with harmful or hazardous drinking, and 15 or more likely indicated alcohol dependence. Consistent condom use was defined as using condoms every time with all sexual partners in the past month.

### Data weighting

Using Stata 17.0, we used a three-stage weighting scheme to achieve population representativeness: (1) Venue weights were the reverse of selection probability (total venues/sampled venues); (2) Patron/worker weights were by venue-specific sampling (total patrons/sampled patrons; venues with <10 workers were assigned weight=1 since they were enumerated entirely); (3) Attendance weights that adjusted for frequency of venue: respondents were assigned monthly “busy days” (Fri-Sun) based on visit frequencies (daily/4–6 times/week=12; 2–3 times/week=10; weekly=4; 2–3 times/month=2.5; ≤monthly=1), with attendance weight = 12 / assigned days. Final weight was the product of these three weights, standardized by dividing by the mean weight to remove selection bias, preserve standard errors, and produce statistical validity.

### Statistical analysis

Our analysis proceeded in five consecutive steps: (1) descriptive characterisation of all social venue-going men; (2) stratification of characteristics by self-reported paid sex during the past 12 months; (3–5) weighted (by PLACE design) adjusted modified Poisson models with robust standard errors examining factors associated with three outcomes—HIV status, virological suppression among HIV+ men, and consistent condom use, with paid sex as the primary exposure in all models. For HIV status, we adjusted for sex payment status, site (fixed effect), age, marital status, education level, and circumcision status as confounding variables.

Variables such as current STI symptoms were excluded from the model as we considered them potential mediators on the causal pathway from paying for sex to HIV acquisition, given that paying for sex plausibly increases STI exposure. However, the directionality between paying for sex and variables such as CMD symptoms, drug use, and hazardous alcohol consumption is less certain. These may either precede or co-occur with the exposure due to shared social or behavioural determinants, therefore, we excluded them from the adjusted models.

For virological suppression, we adjusted for sex payment status, site, age, marital status, and education level, but not AUDIT/CMD/drug use as potential adherence mediators and STI symptoms/condom use as biologically unrelated. For condom use, we adjusted for sex payment status, site, age, marital status, education level, and circumcision status. We excluded HIV status, STI symptoms, drug use, CMD, and AUDIT because they might mediate the relationship between paying for sex and condom use or be influenced by paying for sex, acknowledging the complex/bidirectional relationships some (e.g., drug use, HIV status) may have with it. This approach allowed us to isolate the relationship between paying for sex and HIV-related outcomes while minimizing overadjustment bias caused by potential mediators in the model. All models used the largest-subgroup reference categories to provide stable baselines, reduce variability, and enhance model stability. We reported Wald test p-values. We used modified Poisson regression models to obtain prevalence ratios, which are more intuitive to interpret than odds ratios.

### Ethics statement

This study was approved by the ethics committees at the Medical Research Council of Zimbabwe (MRCZ; MRCZ/A/2867) and the Research Council of Zimbabwe (RCZ). Participants provided informed consent by marking an “X” to confirm that the study’s nature, purpose, benefits, and risks were explained, they received HIV counselling, had time for questions, agreed to testing and discussions, and understood confidentiality. Only the study staff and a witness signed and dated the form; participants did not sign themselves.

## Results

Table 1 presents the sociodemographic, health, and behavioural characteristics of all social venue-going men. The largest proportion of participants was from Harare (62.7%), with the smallest from Gweru (11.9%). Of the men, 39.5% were aged under 30 years. Most had at least a secondary education (90.7%), and more than half were married or living with a partner (52.0%).

**Table 1:** Socio-demographic, health and behavioural characteristics of all social venue- going men – (N=2827)

| **Variable** | **n** | **Weighted column %** |
| --- | --- | --- |
| **Survey site** |  |  |
| Bulawayo | 791 | 13.7 |
| Gweru | 475 | 11.9 |
| Beitbridge | 423 | 12.2 |
| Harare | 1138 | 62.7 |
| **Age** |  |  |
| < 30 years (ref) | 1119 | 39.5 |
| 30 to 39 years | 918 | 32.9 |
| >/= 40 years | 790 | 27.6 |
| **Marital status** |  |  |
| Married/Living with Partner | 1379 | 52.0 |
| Separated/divorced/widowed | 462 | 15.5 |
| Never married | 986 | 32.5 |
| **Education** |  |  |
| None or primary level | 298 | 9.3 |
| Secondary level | 2529 | 90.7 |
| **Non-prescriptive drug use** |  |  |
| Taken any one of the non-prescriptive drugs | 1904 | 68.3 |
| Never took any non-prescriptive drugs | 923 | 31.7 |
| **SSQ-14 mental health** |  |  |
| No symptoms of common mental disorders | 2396 | 71.8 |
| Symptoms suggestive of common mental disorders | 431 | 28.2 |
| **Alcohol use (AUDIT)** |  |  |
| No harmful drinking | 1533 | 57.8 |
| Hazardous or harmful drinking | 535 | 15.2 |
| Alcohol dependence | 759 | 27.0 |
| **Circumcised\*** |  |  |
| No | 1775 | 62.6 |
| Yes | 1033 | 37.4 |
| **Sex payment status** |  |  |
| Paid for sex | 984 | 40.2 |
| Did not pay for sex | 1843 | 59.8 |
| **STI symptoms\*** |  |  |
| Yes | 217 | 12.8 |
| No | 2585 | 87.2 |
| **HIV status\*** |  |  |
| Negative | 2475 | 89.3 |
| Positive | 332 | 10.7 |
| **Condom use** |  |  |
| Consistent | 531 | 15.1 |
| Inconsistent | 2295 | 84.9 |
| **Virological suppression among men living with HIV** |  |  |
| Unsuppressed | 78 | 32.1 |
| Suppressed | 251 | 67.9 |

\* missing information

Behaviourally, 28.2% of respondents tested positive for signs of common mental disorders, 68.3% reported using non-prescription drugs, and 27.0% were alcohol dependent, with nearly 42.0% reporting hazardous or dependent alcohol use. The majority of the men (62.6%) were not circumcised, and 40.1% reported paying for sex in the previous 12 months. Clinically, 10.7% of the men were living with HIV, and among these men, 67.9% had achieved viral suppression. Inconsistent condom use was reported by 84.9% of men. A total of 38 out of 190 venues (20.0%) had at least half of the participants self-reporting as having paid for sex in the past 12 months.

Table 2 presents the distribution of venue-going men who self-reported paying for sex in the past 12 months, stratified by sociodemographic, health, and behavioural characteristics. The weighted percentage of men who self-reported paying for sex was similar in all sites (37.3% - 44.5%) except in Bulawayo (20.0%), where it was lower. Men aged 30–39 years (44.8%) and those who were separated, divorced, or widowed (61.8%) were more likely to report paying for sex compared to other age or marital groups. Paid sex was more common among people with only primary or no education (55.2%) than among those with secondary education (36.8%). Men who reported alcohol dependence (59.7%), had STI symptoms (55.7%), or had symptoms of CMD (58.7%) were likewise more likely to pay for sex. Regarding HIV status, 44.8% of men with HIV and 44.4% of those with suppressed viral load reported paying for sex. Among those who used condoms consistently, 37.2% paid for sex.

**Table 2:** Distribution of venue-going men who self-reported paying for sex in the past 12 months, stratified by sociodemographic, health, and behavioural characteristics (N=2827)s

| **Variable** | **Total** | **n** | **Weighted row (%)** |
| --- | --- | --- | --- |
| **Survey site** |  |  |  |
| Bulawayo | 791 | 162 | 20.0 |
| Gweru | 475 | 162 | 37.3 |
| Beitbridge | 423 | 201 | 43.6 |
| Harare | 1138 | 459 | 44.5 |
| **Age** |  |  |  |
| < 30 years (ref) | 1119 | 383 | 38.1 |
| 30 to 39 years | 918 | 358 | 44.8 |
| > / = 40 years | 790 | 243 | 37.6 |
| **Marital status** |  |  |  |
| Married/Living with Partner | 1379 | 417 | 36.1 |
| Separated/divorced/widowed | 462 | 234 | 61.8 |
| Never married | 986 | 333 | 36.5 |
| **Education** |  |  |  |
| None or primary level | 298 | 118 | 55.2 |
| Secondary level | 2529 | 866 | 36.8 |
| **Non-prescriptive drug use** |  |  |  |
| Taken any one of the non-prescriptive drugs | 1904 | 568 | 35.7 |
| Never took any non-prescriptive drugs | 923 | 416 | 49.8 |
| **SSQ-14 mental health** |  |  |  |
| No symptoms of common mental disorders | 2396 | 797 | 32.9 |
| Symptoms suggestive of common mental disorders | 431 | 187 | 58.7 |
| **Alcohol use (AUDIT)** |  |  |  |
| No harmful drinking | 1533 | 418 | 30.9 |
| Hazardous or harmful drinking | 535 | 225 | 41.0 |
| Alcohol dependence | 759 | 341 | 59.7 |
| **Circumcised\*** |  |  |  |
| No | 1775 | 625 | 42.1 |
| Yes | 1033 | 356 | 37.4 |
| **STI symptoms\*** |  |  |  |
| Yes | 217 | 108 | 55.7 |
| No | 2585 | 869 | 38.0 |
| **HIV status\*** |  |  |  |
| Negative | 2475 | 835 | 39.6 |
| Positive | 332 | 143 | 44.8 |
| **Condom use** |  |  |  |
| Consistent | 531 | 168 | 37.2 |
| Inconsistent | 2295 | 816 | 40.7 |
| **Virological suppression** |  |  |  |
| Unsuppressed | 78 | 35 | 46.7 |
| Suppressed | 251 | 109 | 44.4 |

*\* missing information*

Factors associated with HIV status are shown in Table 3. Notably, paying for sex in the past 12 months was not significantly associated with HIV status (adjusted prevalence ratio [aPR]=0.89, 95% CI: 0.52–1.55), suggesting similar HIV risk between men who paid and those who did not. The analysis revealed that age was a strong predictor of HIV status, with significantly lower prevalence among men under 30 years (aPR=0.22, 95% CI: 0.10–0.47) and marginally lower prevalence among those aged 30–39 (aPR=0.52, 95% CI: 0.27–1.00), compared to those aged ≥40 years. Marital status was also significantly associated with HIV status: married men were less likely to be living with HIV compared to separated/divorced/widowed men (aPR=0.54, 95% CI: 0.32–0.90), while there was no significant difference for never-married men. Circumcision was significantly protective, with circumcised men being less likely to be living with HIV (aPR=0.42, 95% CI: 0.25–0.71).

**Table 3:** Factors associated with HIV status among men tested for HIV at venues (N=2807)

| **Explanatory variables** | **Number**<br>**(n)** | **Number of social**<br>**venue-going men**<br>**who are living with**<br>**HIV: N=332** | **Unweighted**<br>**row %** | **PLACE-**<br>**design**<br>**weighted**<br>**row %** | **Unadjusted**<br>**Prevalence**<br>**Ratios (95%**<br>**CI)** | **Adjusted**<br>**Prevalence**<br>**Ratios (95%**<br>**CI)** | **Adjusted**<br>**Wald**<br>**test P-**<br>**value** |
| --- | --- | --- | --- | --- | --- | --- | --- |
| **Sex payment status in the past 12**<br>**months** |  |  |  |  |  |  |  |
| Paid for sex (ref) | 978 | 143 | 14.6 | 11.9 | 1 | 1 | 0.693 |
| Did not pay for sex | 1829 | 189 | 10.3 | 9.9 | 0.83 (0.48-1.43) | 0.89 (0.52-1.55) |  |
| **Survey sites** |  |  |  |  |  |  |  |
| Bulawayo | 791 | 112 | 14.2 | 11.9 | 0.72 (0.50-1.05) | 0.83 (0.56-1.21) |  |
| Gweru | 475 | 44 | 9.3 | 10.7 | 0.65 (0.42-1.02) | 0.80 (0.52-1.25) |  |
| Beitbridge (ref) | 423 | 69 | 16.3 | 16.5 | 1 | 1 | 0.454 |
| Harare | 1118 | 107 | 9.6 | 9.3 | 0.56 (0.32-0.98) | 0.67 (0.40-1.13) |  |
| **Age** |  |  |  |  |  |  |  |
| < 30 years | 1109 | 34 | 3.1 | 4.5 | 0.22 (0.09-0.56) | 0.22 (0.10-0.47) |  |
| 30 to 39 years | 912 | 101 | 11.1 | 10.3 | 0.51 (0.28-0.91) | 0.52 (0.27-1.00) |  |
| >/= 40 years (ref) | 786 | 197 | 25.1 | 20.1 | 1 | 1 | **<0.001** |
| **Marital status** |  |  |  |  |  |  |  |
| Married/living with a partner | 1369 | 171 | 12.5 | 10.5 | 0.52 (0.29-0.93) | 0.54 (0.32-0.90) |  |
| Separated/divorced/widowed (ref) | 460 | 105 | 22.8 | 20.2 | 1 | 1 | **0.047** |
| Never married | 978 | 56 | 5.7 | 6.5 | 0.32 (0.14-0.77) | 0.86 (0.46-1.59) |  |
| **Level of education** |  |  |  |  |  |  |  |
| None or primary (ref) | 296 | 54 | 18.2 | 17.4 | 1 | 1 | 0.125 |
| Secondary or higher | 2511 | 278 | 11.1 | 10.0 | 0.58 (0.34-0.96) | 0.68 (0.42-1.11) |  |
| **Circumcision status\*** |  |  |  |  |  |  |  |
| Uncircumcised (ref) | 216 | 41 | 19.0 | 23.9 | 1 | 1 | **0.001** |
| Circumcised | 2566 | 286 | 11.2 | 8.7 | 0.33 (0.21-0.51) | 0.42 (0.25-0.71) |  |

*Adjusted for site, age, marital status, education level, sex payment status, and circumcision status*
*p-value is from the Wald test,*
\* *missing information*

Factors associated with virological suppression are shown in Table 4. Paying for sex in the past 12 months was not significantly associated with virological suppression (aPR=1.03, 95% CI: 0.75–1.42). Among 251 men living with HIV, the highest viral suppression levels were in Bulawayo (80.4%), while Harare had the lowest (59.8%, aPR=0.76, 95% CI: 0.52–1.11), though this difference was not statistically significant. Suppression did not significantly vary by age, with men aged ≥40 years having no different suppression from those <30 years (aPR=1.02, 95% CI: 0.66–1.56). Marital status, education, and payment status for sex also did not have any significant associations with virological suppression. No predictors of virological suppression were identified in the adjusted models overall.

**Table 4:** Factors associated with virological suppression among HIV-positive men attending social venues - (N=329)

| **Explanatory variables** | **Number (n)** | **Number of social venue-going men who are virologically suppressed: N=251** | **Unweighted row %** | **PLACE-design weighted row %** | **Unadjusted Prevalence Ratios (95% CI)** | **Adjusted Prevalence Ratios (95% CI)** | **Adjusted Wald test P-value** |
| --- | --- | --- | --- | --- | --- | --- | --- |
| **Sex payment status in the past 12 months** |  |  |  |  |  |  |  |
| Paid for sex | 144 | 109 | 75.7 | 66.7 | 0.97 (0.61-1.53) | 0.97 (0.71-1.34) |  |
| Did not pay for sex (ref) | 185 | 142 | 76.8 | 68.9 | 1 | 1 | 0.864 |
| **Survey site** |  |  |  |  |  |  |  |
| Bulawayo (ref) | 109 | 90 | 82.6 | 80.4 | 1 | 1 | 0.518 |
| Gweru | 44 | 35 | 79.6 | 79.2 | 0.98 (0.78-1.24) | 0.94 (0.75-1.18) |  |
| Beitbridge | 69 | 49 | 71.0 | 73.8 | 0.92 (0.74-1.13) | 0.90 (0.69-1.17) |  |
| Harare | 107 | 77 | 72.0 | 59.8 | 0.74 (0.47-1.17) | 0.76 (0.52-1.11) |  |
| **Age** |  |  |  |  |  |  |  |
| < 30 years (ref) | 35 | 20 | 57.1 | 76.8 | 1 | 1 | 0.347 |
| 30 to 39 years | 98 | 71 | 72.5 | 51.8 | 0.67 (0.37-1.22) | 0.71 (0.41-1.21) |  |
| ≥ 40 years | 196 | 160 | 81.6 | 74.5 | 0.97 (0.67-1.46) | 1.02 (0.66-1.56) |  |
| **Marital status** |  |  |  |  |  |  |  |
| Married/living together | 167 | 134 | 80.2 | 71.0 | 0.93 (0.62-1.40) | 0.98 (0.68-1.40) |  |
| Separated/widowed/divorced | 107 | 81 | 75.7 | 57.5 | 0.76 (0.43-1.34) | 0.81 (0.48-1.37) |  |
| Never married | 55 | 36 | 65.5 | 76.0 | 1 | 1 | 0.683 |
| **Level of education** |  |  |  |  |  |  |  |
| None or primary level (ref) | 53 | 38 | 71.7 | 78.0 | 1 | 1 | 0.314 |
| Secondary or higher | 276 | 213 | 77.2 | 66.2 | 0.85 (0.62-1.17) | 0.88 (0.68-1.13) |  |

*Adjusted for site, age, marital status, education level, and sex payment status*
*All percentages presented in this table are weighted using PLACE-design weights to account for differential selection probabilities across sites and strata.*
*p-value is from the Wald test*

Factors associated with reporting consistent condom use in the previous month are shown in Table 5. Our findings demonstrate that consistent condom use in the previous month among men who frequented social venues was comparatively low across all sites. In adjusted analysis, notably, paying for sex in the past 12 months was not significantly associated with consistent condom use (aPR=0.87, 95% CI: 0.57–1.34). Men in Harare were significantly less likely than those in Beitbridge to use condoms consistently (aPR=0.58, 95% CI: 0.39–0.87). Marital status was strongly associated with condom use: married men were considerably less likely to use condoms consistently than never-married men (aPR=0.28, 95% CI: 0.19–0.42). There was weak evidence that men with no or primary education level (aPR=0.71, 95% CI: 0.48–1.04) were less likely to report consistent condom use compared with men with secondary or higher education level.

**Table 5:** Factors associated with consistent condom use among all venue-going men - (N=2,827)

| **Explanatory variables** | **Number**<br>**(n)** | **Number of social**<br>**venue-going men who**<br>**self-reported**<br>**consistent condom**<br>**use in the past month:**<br>**N=561** | **Unweighted**<br>**row %** | **PLACE-**<br>**design**<br>**weighted**<br>**row %** | **Unadjusted**<br>**Prevalence**<br>**Ratios (95%**<br>**CI)** | **Adjusted**<br>**Prevalence**<br>**Ratios (95%**<br>**CI)** | **Adjusted Wald**<br>**test P-**<br>**value** |
| --- | --- | --- | --- | --- | --- | --- | --- |
| **Sex payment status in the past 12**<br>**months** |  |  |  |  |  |  |  |
| Paid for sex | 984 | 168 | 17.1 | 14.0 | 0.88 (0.60-1.31) | 0.87 (0.57-1.34) | 0.528 |
| Did not pay for sex (ref) | 1843 | 363 | 19.7 | 15.8 | 1 | 1 |  |
| **Survey sites** |  |  |  |  |  |  |  |
| Bulawayo | 791 | 167 | 21.1 | 18.7 | 0.91 (0.69-1.21) | 0.72 (0.51-1.00) |  |
| Gweru | 475 | 103 | 12.7 | 18.4 | 0.90 (0.66-1.21) | 0.84 (0.62-1.13) |  |
| Beitbridge (ref) | 423 | 88 | 20.8 | 20.6 | 1 | 1 | **0.041** |
| Harare | 1138 | 173 | 15.2 | 12.5 | 0.61 (0.41-0.91) | 0.58 (0.39-0.87) |  |
| **Age** |  |  |  |  |  |  |  |
| < 30 years (ref) | 1119 | 283 | 25.3 | 20.5 | 1 | 1 | 0.512 |
| 30 to 39 years | 918 | 133 | 14.5 | 10.7 | 0.52 (0.36-0.75) | 0.94 (0.69-1.29) |  |
| > / = 40 years | 790 | 115 | 14.6 | 12.5 | 0.61 (0.36-1.04) | 1.30 (0.79-2.14) |  |
| **Marital status** |  |  |  |  |  |  |  |
| Married/living with a partner | 1379 | 104 | 7.5 | 7.7 | 0.30 (0.18-0.50) | 0.28 (0.19-0.42) |  |
| Separated/divorced/widowed | 462 | 130 | 28.1 | 18.0 | 0.71 (0.48-1.04) | 0.69 (0.45-1.06) |  |
| Never married (ref) | 986 | 279 | 30.2 | 25.4 | 1 | 1 | **<0.001** |
| **Level of education** |  |  |  |  |  |  |  |
| None or primary | 298 | 48 | 16.1 | 11.0 | 0.71 (0.45-1.12) | 0.71 (0.48-1.04) |  |
| Secondary or higher (ref) | 2529 | 483 | 19.1 | 15.5 | 1 | 1 | **0.080** |
| **Circumcision status\*** |  |  |  |  |  |  |  |
| Uncircumcised | 1775 | 311 | 17.5 | 14.2 | 0.86 (0.59-1.27) | 1.06 (0.76-1.48) |  |
| Circumcised (ref) | 1033 | 217 | 21.0 | 16.5 | 1 | 1 | 0.733 |

*Adjusted for site, age, marital status, education level, circumcision status, and sex payment status*
*p-value is from the Wald test*
\* *missing information*

## Discussion

To better inform prevention and treatment programming among men in Zimbabwe, we assessed HIV prevalence, virological suppression, and HIV-related behaviours among men at social venues in four cities and towns. Among 2,827 venue-going PLACE men, 984 (40.1%) reported paying for sex in the previous year. HIV prevalence was 10.7% overall, and among these men living with HIV, only 68.9% were virologically suppressed; below the UNAIDS 2030 target of 86%(1). Few men (15.1%) reported consistent condom use. We found no significant differences in HIV status, virological suppression, or consistent condom use between men who paid for sex and those who did not. We highlighted substantial risk and high HIV prevention and treatment needs among all venue attendees.

Our venue-based surveys effectively reached men who pay for sex, a group often missed in traditional surveys. We argue that conducting surveys during peak times improved efficiency, engaging local stakeholders fostered community acceptance, and probability sampling enhanced representativeness and generalizability to venue-going men. However, our study did have a number of limitations. Self-reported data could have resulted in social desirability/recall bias(34–38). The cross-sectional design could prevent causal/temporal inferences, and demographic/cultural/socioeconomic/healthcare variations, and non-random province selection may constrain the generalizability of our results. In addition, our small virological suppression sample and our inability to distinguish condom use by partner type (spouse vs. others) for married men limit robustness. Future research should differentiate condom use by partner.

We applied sampling weights in PLACE surveys. Larger discrepancies between weighted and unweighted estimates occurred for venue-clustered variables (age, education, drug use, paid sex status), where weighting amplifies/reduces the influence of venues with distinct characteristics. Paid sex showed strong behavioural clustering: 20.0% of venues contained >50.0% of men reporting it, with methodological (weighting), epidemiological (hotspot targeting), and programmatic (resource allocation) implications. Subgroups over-/under-sampled relative to the broader population received heavier/lighter weights, while evenly distributed variables required smaller adjustments.

Among Zimbabwean social venue attendees, 40.1% reported paying for sex in the past year (with substantial site variation), exceeding rates from East African venue samples (20.0–22.5%, likely reflecting underreporting)(39), SSA population-representative surveys (2.7% past year; 8.0% lifetime)(40), and Zimbabwe DHS 2015 (4.0% past year; 18.0% lifetime—probable underestimates)(4). The high rate of recent paid sex (49.0% in past 4 weeks) confirms ongoing high-risk exposure. While direct African venue data are scarce, studies of high-risk occupational groups (e.g., truck drivers/miners: 6.6–47.8% past year) show comparable prevalences. Many individuals from these occupational groups likely visit the venues included in the study, making them a relevant comparison group(41–45). However, other studies of high-risk subgroups report lower proportions(46–50).

HIV prevalence among venue-going men aged ≥16 years was 10.7% in our study, with no significant difference by sex payment status. This aligns with Zimbabwe national surveys: ZIMPHIA 2020 (10.2% men 15-49 years)(51), and ZDHS 2015 (10.5% men ≥15 years)(4). All three sources show higher prevalence among men ≥35 years. Notably, ZDHS 2015 found men reporting paid sex were more likely to be LHIV than non-payers. While contextual differences limit direct comparison, our prevalence was lower than among South African venue men (15.5%)(10). These findings confirm that venue-going men constitute a significant proportion of people LHIV, necessitating targeted interventions.

Virological suppression among men LHIV was lower in our study (68.9%) than in ZIMPHIA 2020 (73.0% among men ≥15 years)(51). This gap may reflect poorer ART engagement in high-risk venue-going populations and contextual/demographic differences. Both rates fall substantially below UNAIDS’ 86.0% suppression target, representing the proportion of all people living with HIV who are virologically suppressed if the targets are met(1).

Consistent condom use was low: only 15.1% overall and 14.0% of recent sex-payers self- reported consistent use in the past month. While inconsistent use is commonly documented across populations(41–44, 48, 52–56), direct comparison with ZDHS 2015 (90.2% condom use at last paid sex among payers)(4) is limited by our measure’s lack of partner-type distinction. Other high-risk groups show similar gaps: 34.8% of truck drivers reported consistent use (past 3 months)(52), and 35.7% used condoms at last vaginal sex in East African venues in past year(29). Despite generally high condom access, these patterns underscore widespread unsafe sex among venue-going men, elevating HIV transmission risk.

Despite venue-going men exhibiting higher overall HIV risk and poorer prevention/care engagement than the general population, we observed no elevated HIV prevalence among those reporting paid sex versus non-payers. This apparent anomaly may stem from underreporting of paid sex by men LHIV due to stigma/social desirability bias or genuine behavioural differences (e.g., reduced sexual activity). This anomaly may also arise due to comparable risk behaviours among non-payers (e.g., multiple/casual partnerships) diminishing observable prevalence differences.

Comprehensive strategies are essential to improve HIV testing, ART adherence, retention, condom use, and PrEP uptake among all venue-going men. Effective approaches include targeted sexual education, counselling, peer outreach, men-focused messaging, routine HIV self- testing, simplified ART regimens, adherence clubs, mHealth interventions, and safe drop-in centres, with venue owner/manager engagementϑ(57–59). Prioritizing treatment as prevention ensures early ART initiation and sustained care for men living with HIV.

Key HIV prevention enhancements include offering oral/injectable PrEP (e.g., CAB-LA) and discreet condom stations at venues, implementing community ART distribution (PODIs) and refill groups (CARGs) to reduce clinic waits/stigma barriers(60, 61), multi-venue kit distribution for convenient testing access(62, 63), and targeted Undetectable=Untransmittable (U=U) campaigns with emotional support to boost ART uptake.

## Conclusion

Our study highlights significant unmet HIV treatment/prevention needs among Zimbabwean venue-going men, regardless of sex-payment history. Venue sampling proved feasible for reaching the diverse, hard-to-reach population of men having sex with FSWs. These findings address the evidence gap on HIV in African venue-going men. Future interventions should explore on-site service delivery at social venues to reduce HIV risk, while longitudinal studies could monitor changes in prevalence, adherence, and sexual behaviours.

## Declarations

Ethics approval and consent to participate were obtained in accordance with the principles outlined in the Declaration of Helsinki, ensuring that the study adhered to ethical standards for research involving human subjects. This study was approved by the Medical Research Council of Zimbabwe (MRCZ/A/2867) and the Research Council of Zimbabwe (RCZ). All participants provided written informed consent

## Consent for publication

Not applicable

## Availability of data and materials

The data that supports the findings of this study are openly available in: Open Science Framework: https://osf.io/rsd9z/files/osfstorage

## Authors’ contributions

GM planned and conducted the analysis, and GM wrote the first draft. STC, FM, JM, MM, TC, SC, LN, and MM led the data collection. EM converted the final versions of the survey tools— including surveys and questionnaires—into electronic formats and designed the database. JRH, EF, and FMC provided critical revisions to the article, particularly the discussion. FM, BR, KG, SW, JRH, and FMC were involved in the conception of the study. STC, BR, SW, JRH, EF, and FMC were involved in the interpretation of results and critical revision of the article. AM, RY, BM, and OM provided technical oversight for program implementation. All authors contributed to the writing and have read and approved the final version.

## Acknowledgements

We would like to thank the survey team and all the participants who took part in the study.

